# Clinically relevant variability of the lipidome in people with type 2 diabetes

**DOI:** 10.64898/2026.07.06.26357365

**Authors:** Simon Maria Kienle, Tommi Suvitaival, Martin Bæk Blond, Joana Mendes Lopes de Melo, Mads Almose Røpke, Karolina Sulek, Joachim Størling, Peter Rossing, Cristina Legido-Quigley

## Abstract

**Background:** Besides hyperglycemia, type 2 diabetes (T2D) is characterized by dyslipidemia, which is typically assessed using traditional clinical lipid measurements. However, molecular plasma lipids beyond these traditional markers can provide additional information about an individual’s health status. For molecular lipids to be used effectively, certain characteristics, such as their temporal variability, need to be determined.

**Methods:** We analyzed the plasma lipidome for three consecutive time points, each three months apart, of 51 individuals with T2D using targeted liquid chromatography coupled to mass spectrometry (LC-MS). 513 lipid species across 25 (sub)classes were quantified by this approach and the temporal variability were calculated. Moreover, to identify sex differences in the plasma lipidome, we analyzed 914 samples of a cross-sectional T2D cohort with the same approach.

**Results:** Neutral lipids and phosphatidylserine had the highest temporal variability which was independent of their platform-specific variability. In contrast, glycosphingolipids were found to be relatively stable over time in individuals with T2D. Acyl-chain analysis revealed generally similar variability in the acyl-chain groups but indicated a higher temporal variability in medium-length acyl-chains. Lipid-sex association analysis showed markedly higher sphingomyelins, phosphatidylcholines, and phosphatidylethanolamines in women and higher acylcarnitines in men. Overall, approximately one-third of measured lipids showed significant sex differences independent of age, BMI, diabetes duration, glycemic control, and medication use.

**Conclusions:** Our findings provide insights into temporal variability of molecular lipids. This variability should be considered when assessing novel lipid biomarkers. Likewise, sex differences in these lipids need to be considered in precision medicine for diabetes management.

**Plain language summary:** The human blood contains many types of fat particles, also referred to as lipids. Currently only a specific subset of these blood lipids is used for decision making in healthcare. However, lipids beyond these traditional clinical lipids can add information about the health status of an individual and thus can support more precise decision making. To measure these non-traditional lipids, modern technologies, like mass spectrometry-based lipidomics, can be applied. However, important prerequisites need to be assessed for both the non-traditional lipids as well as the used technologies to evaluate their usability in healthcare. We determined some of these prerequisites for blood samples from individuals living with type 2 diabetes and identified differences in many lipid levels between men and women.

## INTRODUCTION

Lipids are an important class of biomolecules involved in many vital cellular processes. Due to their hydrophobic nature, lipids are the major constituents of biological membranes^1,2^. Moreover, they serve as storage molecules for surplus energy and participate in cellular signaling processes^1,2^.

From a chemical point of view, lipids are a heterogeneous group of compounds containing fatty acyl/alkyl/alkenyl, sphingosine, or isoprene moieties as their hydrophobic building blocks^3,4^. Due to the biological importance of lipids, imbalances in their homeostasis contribute to the development and progression of serious conditions, such as chronic inflammation, cardiovascular disease (CVD), diabetes, and neurodegenerative diseases^5–8^. Hence, it is important to comprehensively profile the level of lipid molecules.

To analyze lipid molecules comprehensively (known as the lipidome), mass spectrometry (MS) is often used since it provides the required sensitivity and specificity^5,7,9^. However, due to the inherent chemical complexity of the lipidome, represented by for example isobaric lipid species and isomeric species, it is challenging to structurally characterize lipids by MS only^3,9^. To accomplish this, liquid chromatographic (LC) separation combined with tandem MS (LC–MS/MS) is typically applied^5,9–11^. In the past decade, targeted LC-MS/MS lipidomics has been shown to allow the profiling of the human lipidome in blood samples in great detail, enabling the investigation of associations between the lipidome and anthropometric factors, diseases or disease stages, as well as supporting biomarker discovery^12–18^.

For the discovery of biomarkers, lipidomic data from thousands of individuals need to be collected, thus lipidomics platforms need to have sufficient stability over the course of analysis and their platform-specific variability needs to be low.

Lipid concentrations are affected by a range of factors and can fluctuate over time^19^. Consequently, the use of lipid biomarkers for the treatment of individual patients requires assessment of their variability to determine thresholds or reference ranges and sampling intervals.

In recent years it has become obvious that biological sex has a strong influence on circulating lipid profiles^6,12–14,19–21^and since some diabetes associated diseases show sex-specific differences^22^, determining the influence of sex on lipidomic data in a (T2D)-specific population is crucial for enabling sex-adapted therapies. Therefore, sex-specific lipid profiles must be accurately assessed and incorporated when examining associations between specific lipids and clinical outcomes such as T2D and CVD.

Here we report the use of a reversed-phase targeted LC-MS/MS lipidomics platform to profile 513 lipid species from human blood samples. This analysis pipeline allowed us to characterize both isobaric and isomeric lipid species, supporting a better understanding of physiological processes and lipid metabolism. After determining the platform-specific variability of the lipidomic analysis, we profiled the plasma lipidome in a cohort of 51 individuals living with T2D at three consecutive time points (each three months apart) to assess their temporal lipid variability, which is fundamental for evaluating the usability of lipid classes and species for future MS-based lipidomic biomarker discovery. Moreover, we measured the lipidomic profile in a cross-sectional cohort with 957 individuals living with T2D, to determine sex-specific lipid differences.

## RESULTS

Due to the diversity of lipids, an important prerequisite for obtaining a representative fingerprint of circulating lipids from an individual is the broad analytical coverage of the lipid classes and the respective lipid molecules^9–11,13^. Recently, high-coverage LC-MS/MS lipidomics methods based on a targeted approach with triple quadrupole (QqQ) instruments have been reported to fulfill this criterion^5,9,11,16^ and thus we established a targeted lipidomics platform (Figure 1). In this platform we detect 513 transitions belonging to 25 lipid (sub)classes via a scheduled/dynamic multiple reaction monitoring (dMRM) approach. To be able to distinguish between isobaric and isomeric lipid molecules we employed C18 reversed-phase LC prior the MS/MS analysis (a complete list of MRM transitions and retention times is shown in the Supplementary).

**Fig. 1.**
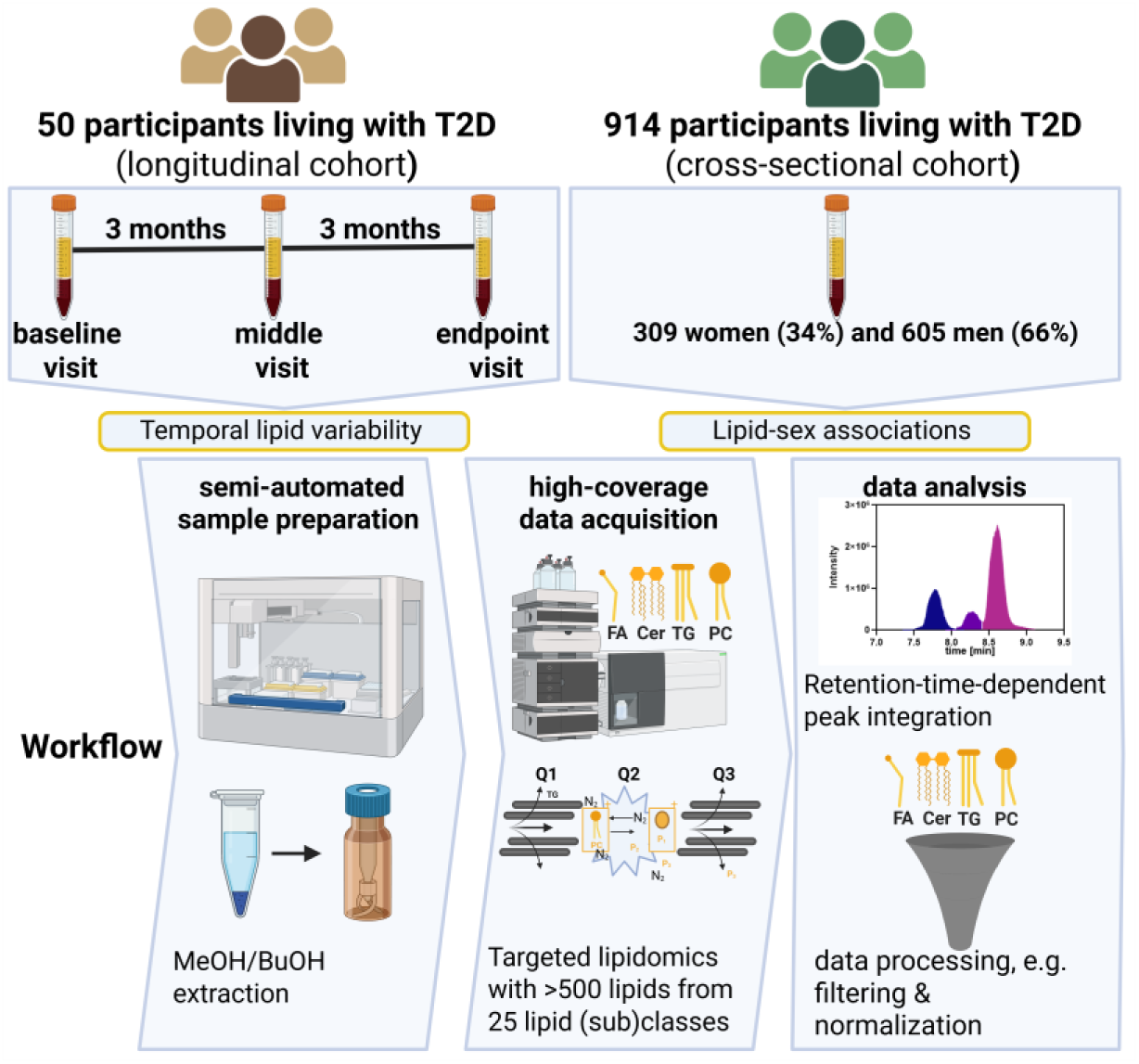
Overview of cohorts and analytical pipeline. Schematic representation of the study design and analytical pipeline. The longitudinal cohort included 50 individuals with type 2 diabetes (T2D) sampled at baseline, middle, and endpoint visits over two consecutive 3-month intervals. The cross-sectional cohort comprised 957 individuals with T2D. The workflow illustrates semi-automated sample preparation using MeOH/ BuOH extraction, targeted lipidomics acquisition covering more than 500 lipid species across 25 lipid (sub)classes, and data processing steps including retention-time-dependent peak integration, filtering, and normalization.

### Linearity and platform-specific lipid variability

To test the quantitative performance for the lipid analytes, we assessed the measurement linearity by spiking exogenous lipids in different concentrations into plasma (Sup Figure 1). For all tested lipid species, we observed linear increases in signal intensity across three orders of magnitude of increasing concentration. However, for the highest and lowest concentration, we observed detector signal saturation or limit of detection, respectively. Since we use a C18 column for lipid separation prior to MS analysis, we additionally investigated sample carry-over between runs. Even though a small number of lipids showed carry-over, its contribution to the signal intensity was minimal, except for the free fatty acid FFA(14:0) and the diacylglycerol DG(16:0_16:0) (Sup Figure 1).

Besides accurate analyte quantification, high reproducibility is also crucial when investigating plasma lipid profiles in clinical settings. Thus, we investigated platform-specific lipid variability by determining the coefficient of variation (CV) for each lipid species (Figure 2). In general, we observed good reproducibility indicated by an average CV of 8.6% across all lipid species based on 30 pooled plasma sample injections over the course of an analysis sequence of over 370 measurements. We did, however, observe heterogeneity in CVs between lipid classes. While all lysoalkylphosphatidylcholine (LPC(O)) lipid species had low CVs (average CV of 3.7% in the lipid class), the cholesteryl esters (CEs) had highest CVs (CV of 23.1%). Overall, lipids with a relatively simple molecular structure had the lowest CVs. In particular, lipids with only one acyl-chain, such as, acylcarnitines (CARs), lysophosphatidylcholines (LPCs) as well as its alkyl- and alkenyl-ether lipids (LPC(O/P)), had low CVs, with the exception of CEs. In summary, the established lipidomics method is well suited for profile plasma lipids showing low platform-specific variability while capturing linear changes of analytes with a minimal carry-over of analytes. All these features are prerequisites for MS applications in clinical settings.

**Fig. 2.**
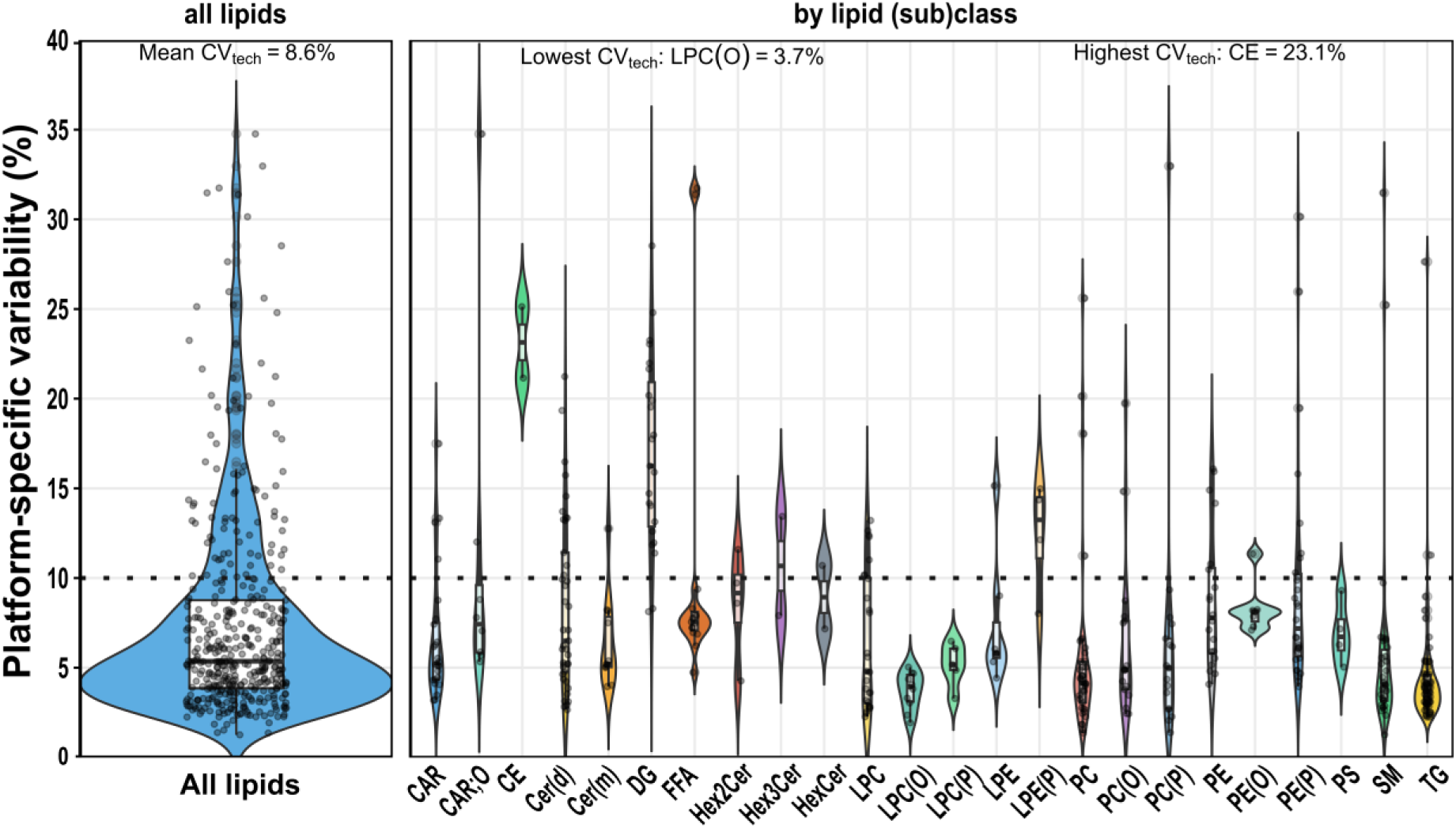
Platform-specific variability of lipid measurements across all lipids and lipid (sub)classes. Violin plots show the distribution of platform-specific variability in % for all individual lipid species and by lipid (sub)classes after data processing for technical replicates. The mean platform-specific variability for all lipids is 8.6%, but lipid classes differ in variability. Lowest variability is found in the lipid class LPC(O) with= 3.7%, while lipids from CE lipid class show in average the highest variability with 23.1%. Boxplots and individual lipid species as black points are show within the violin plots. Dotted line indicates 10% CV. CAR, acylcarnitines, CAR;O, hydroxyl-acylcarnitines CE, cholesteryl ester; Cer(d), ceramide; Cer(m), deoxyceramide; DG, diacylglycerol; HexCer, monohexosylceramide; Hex2Cer, dihexosylceramide; Hex3Cer, trihexosylceramide; LPC, lysophosphatidylcholine; LPC(O), lysoalkylphosphatidylcholine; LPC(P), lysoalkenylphosphatidylcholine; LPE, lysophosphatidylethanolamine; LPE(P), lysoalkenylphosphatidylethanolamine; PC, phosphatidylcholine; PC(O), alkylphosphatidylcholine; PC(P), alkenylphosphatidylcholine; PE, phosphatidylethanolamine; PE(O), alkylphosphatidylethanolamine; PE(P), alkenylphosphatidylethanolamine; SM, sphingomyelin; TG, triacylglycerol; Data table with values for each lipid species is provided in supplemental source Table 4.

### Plasma lipid variability over time in people living with diabetes

There are various internal and external factors affecting the plasma lipid profile. Understanding the intra- and inter-individual lipid variability is essential for the generation and interpretation of lipidomics results, especially when considering the use of lipid biomarkers in clinical settings^6,19^. To determine the variability in plasma lipid levels in people with T2D, we measured the plasma lipidome in a longitudinal cohort of 51 individuals at three consecutive time points, each three months apart (for clinical characteristics see Table S1).

From the 513 measured lipids, 445 passed the data filtering steps, including 97 neutral lipids (TGs, DGs, and CEs), 150 glycerophospholipids (PCs, PC(P)s, PC(O)s, PEs, PE(P)s, PE(O)s, PSs), 57 lysoglycerophospholipids (LPCs, LPC(P)s, LPC(O)s, LPEs, LPE(P)s, LPE(O)s), 94 sphingolipids (Cers, SMs, HexCers), 15 free fatty acids (FFAs) and 32 acylcarnitines (CARs). For each lipid (sub)class, we quantified the within-individual temporal lipid variability to evaluate class-specific fluctuations during the study period (Figure 3). Phosphatidylserine (PS) showed the highest temporal variability (mean 59%), followed by glycerolipids (TG and DG). In contrast, glycosphingolipids (Hex3Cers, Hex2Cers, and HexCers) exhibited the lowest degree of fluctuation, indicating that these lipid classes have stable levels in human plasma, which is an important prerequisite for biomarker application.

**Fig. 3:**
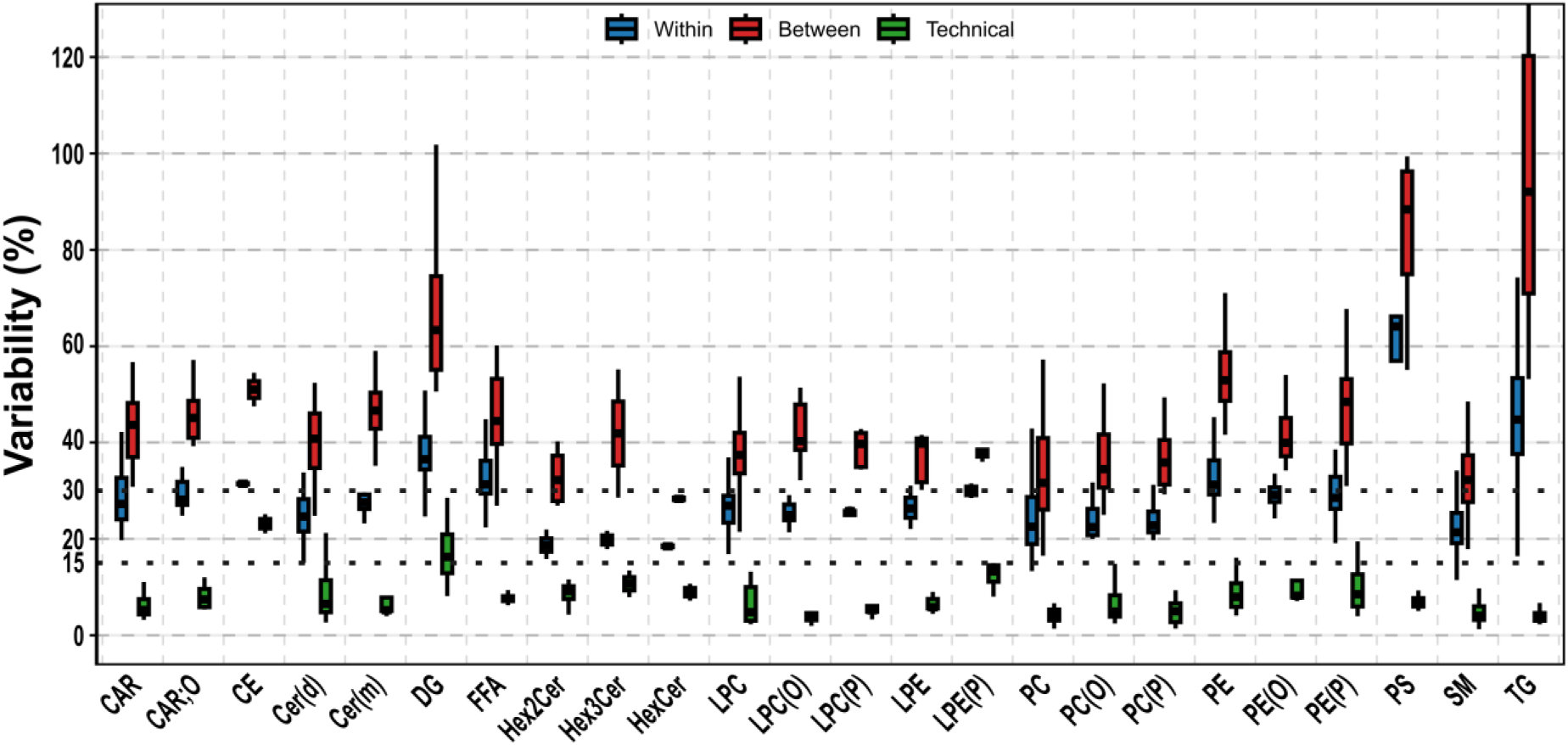
Temporal within-individual and between-individual variability in lipid levels. Within-individual and between-individual variability is high in PS, TG, and DG lipid classes. Variability in percent (%) is shown for each lipid (sub)class, partitioned into temporal within-individual (blue), between-individual (red), and technical (green) components. While technical variability remained consistently low across subclasses, PS and neutral lipid (TG and DG) have both, a high temporal variability within an individual and a high between-individual variability. Box plots display the distribution of variablility estimates across detected lipid species within each subclass. Dotted lines indicate 15% and 30% vriability. CAR, acylcarnitines, CAR;O, hydroxyl-acylcarnitines CE, cholesteryl ester; Cer(d), ceramide; Cer(m), deoxyceramide; DG, diacylglycerol; HexCer, monohexosylceramide; Hex2Cer, dihexosylceramide; Hex3Cer, trihexosylceramide; LPC, lysophosphatidylcholine; LPC(O), lysoalkylphosphatidylcholine; LPC(P), lysoalkenylphosphatidylcholine; LPE, lysophosphatidylethanolamine; LPE(P), lysoalkenylphosphatidylethanolamine; PC, phosphatidylcholine; PC(O), alkylphosphatidylcholine; PC(P), alkenylphosphatidylcholine; PE, phosphatidylethanolamine; PE(O), alkylphosphatidylethanolamine; PE(P), alkenylphosphatidylethanolamine; SM, sphingomyelin; TG, triacylglycerol. Data table with values for each lipid species is provided in supplemental source Table 4

Yet, temporal variability of molecular lipid levels in human plasma might not be identical for molecular species within a lipid class, but might differ according to properties, such as, the acyl-chain length and the degree of unsaturation. Thus, we analyzed the data by grouping lipids according to their acyl-chain(s) into lipids with saturated (SFA), monounsaturated (MUFA) and polyunsaturated fatty acids (PUFA). Moreover, besides grouping by saturation, we additionally clustered lipid acyl-chain(s) into biologically relevant medium-(C6-12), long-(C13-21) and very-long chain-lengths (>C21) as well as into even and odd chain lengths. We found no statistically significant difference in the temporal variability for lipids with SFAs, MUFAs and PUFAs (Sup Figure 2) and neither a statistically significant difference in variability between lipids with odd and even acyl-chains, nor between medium-, long and very-long acyl-chain(s) (Sup Figure 2). This indicates that the above-mentioned temporal variability effects are lipid class specific.

To investigate which lipid levels per lipid classes vary most between participants, we next analyzed the between-individual variability for each lipid (sub)class (Figure 3). Again, the highest between-individual variability was observed for TGs and PSs, as well as DGs. In contrast, sphingolipid, including SMs, HexCers and Hex2Cers, had the lowest variability indicating that these lipid classes are rather stable in their plasma levels also between individuals. With respect to the acyl-chain(s) analysis, again neither a statistically significant difference was observed between SFAs, MUFAs and PUFAs, nor in acyl chain-length or -parity (Sup Figure 3).

For biomarkers application it is important that the between-individual variability is greater than the within-individual variability. In line with this, on average, all lipid classes exhibited greater variability between individuals than the temporal variability within individuals. To further quantify this, we calculated the ratio of the between-individual and within-individual variability (Table S2). In general, TGs, DGs, and glycosphingolipids (Hex2Cers and Hex3Cers) had the highest ratios and thus vary most between individuals but less within an individual. On the lipid species level, TG(18:1_40:1) emerged with the highest ratio, showing low longitudinal variability but high between-individual variability. In other words, the level of TG(18:1_40:1) appears to be characteristic of a given individual and stable over time in the studied cohort. The measured temporal lipid variability is a product of level variations caused by lipid metabolism changes, diet and environmental factors as well as platform-specific variability. In our case we found that the temporal variability in CEs is almost as high as the platform-specific variability whereas this is low for PS and TG (Figure 3).

In summary, we determined temporal stability of lipids in human plasma which is a critical parameter for clinical biomarker usage. Moreover, we found that in TGs and DGs as well as PSs between-individual variability dominate temporal variability.

### Plasma lipid association to sex in 914 individuals living with type 2 diabetes

In recent years, it became evident that variations in treatment responses and disease progression are related to biological differences between men and women^19,22^. To examine sex-dependent lipidomic profiles, we extended our analysis to a larger, cross-sectional, T2D cohort; we determined plasma lipid profiles from the Thousand&2 cohort, which consists of 605 (66,2%) men and 309 women (N = 914 individuals in total; Table 1)^23^. From the 513 measured lipids, 455 passed quality control, including 95 glycerolipids, 216 glycerophospholipids, 94 sphingolipids, 3 sterol lipids, and 47 fatty acyls. At the lipid class level, 13 out of 25 (sub)classes were significantly associated with sex after adjusting for age (Figure 4a and Supplementary Table S2). The lipid classes most strongly influenced by sex included sphingomyelin (SM), phosphatidylethanolamine (PE) and phosphatidylcholine (PC), including the alkyl-(PC(O); PE(O)) and the alkenyl-ether (PC(P); PE(P)) subclasses of both, as well as glycosphingolipids (HexCer; Hex3Cer) and FFA. All these lipid (sub)classes were higher in their levels in women. The lipid (sub)classes showing a significantly higher level in men were acylcarnitines (CAR) and hydroxylated acylcarnitines (CAR;O). Most of these lipid class-specific associations to sex were additionally validated in the longitudinal cohort of 51 people with T2D (Sup Figure 4).

**Fig. 4:**
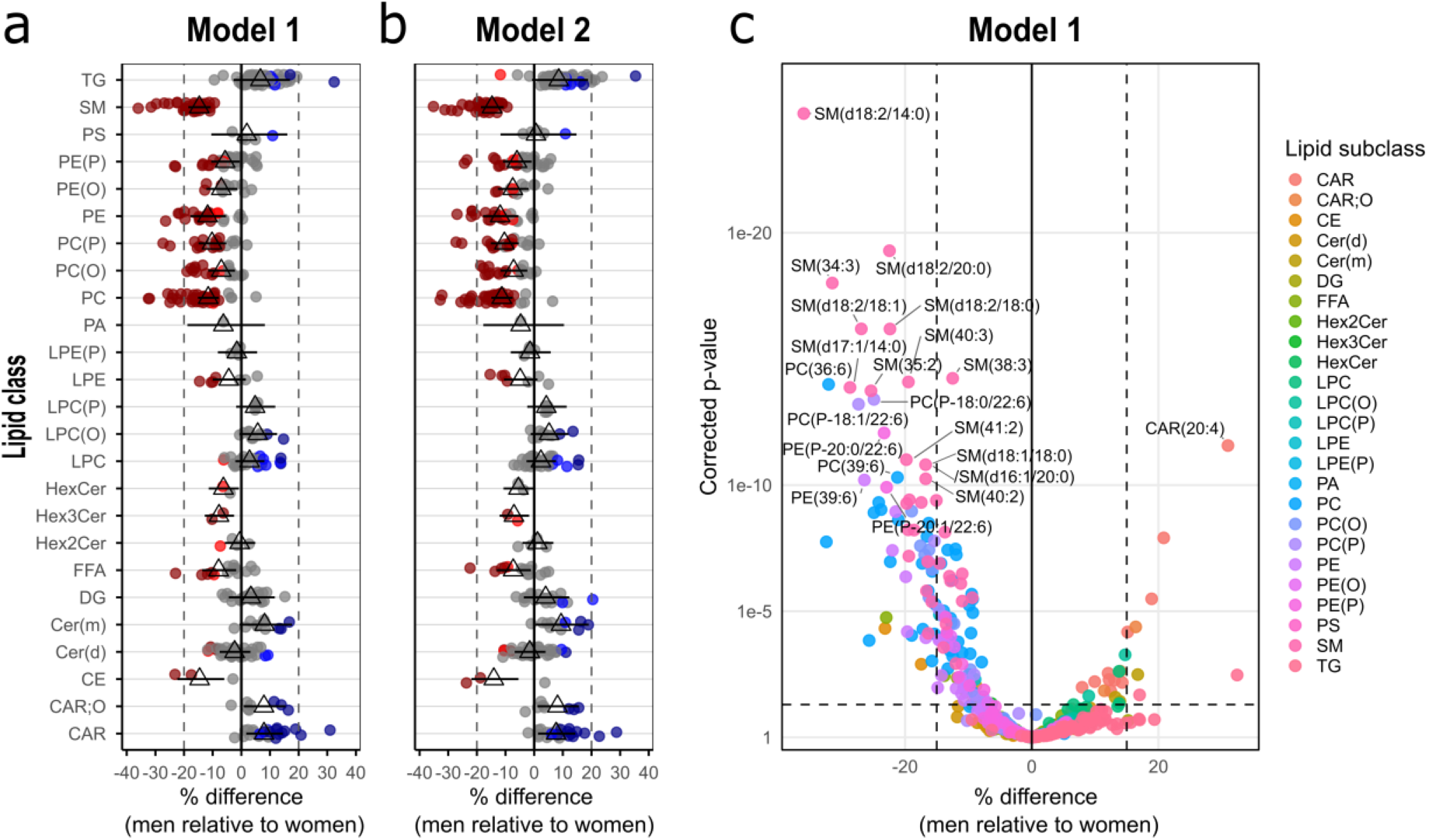
Associations between sex and the levels of lipid subclasses and species. **(a-b):** SM, FFAs, HexCers, Hex3Cers, CE, PC and PE, as well as subclasses thereof, are higher in women in both models used. Linear regression analysis between sex and log-transformed concentrations of each lipid species was performed adjusting for age ((a) Model 1), and for age, BMI, diabetes duration, smoking, HbA1c, statin-, metformin-, DPP4-, liraglutide-, sulfonylurea-treatment ((b) Model 2) for 957 individuals living with T2D. The beta coefficients were converted to percentage difference for men relative to women. Light and dark red indicates p-value <0.05 and FDR-corrected p-values <0.05, respectively for lipids that a lower in men as in women. Light and dark blue indicates p-value <0.05 and FDR-adjusted p-values <0.05, respectively for lipids that a higher in men as in women. Gray points indicate non-statistically significant associated lipids. Triangle and whiskers represent mean and 95% confidence intervals for the corresponding lipid classes, respectively. **(c)** Associations between sex and lipid species plotted by their percent differences and FDR-corrected p-values. Top lipid species are indicated with their name and circles are colored according to the lipid (sub)classes. Data table with values for each lipid species is provided in supplemental source Table 2 and 3.

**Table 1:** Thousand&2 clinical characteristics by sex. Data for continuous variables are presented as median [IQR] and categorical variables as number of individuals and as percentage in brackets. HbA1c: hemoglobin A1c, HDL: high-density lipoprotein, LDL: low-density lipoprotein.

| Variable<sup>1</sup> | All<br>N = 914<sup>1</sup> | Men<br>N = 605<sup>1</sup> | Women<br>N = 309<sup>1</sup> |
| --- | --- | --- | --- |
| **Age (years)** | 65.6 [59.2, 71.7] | 65.7 [59.4, 70.9] | 65.1 [58.2, 72.4] |
| **Diabetes duration (years)** | 11.5 [5.5, 17.0] | 12.0 [6.0, 17.0] | 10.0 [5.0, 17.0] |
| **Body mass index (kg/m<sup>2</sup>)** | 29.3 [26.3, 33.3] | 29.3 [26.6, 33.0] | 29.4 [25.9, 34.1] |
| **Systolic blood pressure (mmHg)** | 133.0 [125.0, 145.0] | 130.0 [121.0, 145.0] | 135.0 [125.0, 150.0] |
| **Diastolic blood pressure (mmHg)** | 80.0 [75.0, 85.0] | 80.0 [74.0, 85.0] | 80.0 [75.0, 85.0] |
| **Smoking status** |  |  |  |
| *Never* | 388 (42%) | 228 (38%) | 160 (52%) |
| *Former* | 393 (43%) | 286 (47%) | 107 (35%) |
| *Current* | 133 (15%) | 91 (15%) | 42 (14%) |
| **Albuminuria category** |  |  |  |
| *Makro* | 63 (7.0%) | 46 (7.7%) | 17 (5.7%) |
| *Mikro* | 146 (16%) | 116 (19%) | 30 (10%) |
| **LDL cholesterol (mmol/L)** | 2.0 [1.6, 2.6] | 1.9 [1.5, 2.5] | 2.1 [1.7, 2.8] |
| **HDL cholesterol (mmol/L)** | 1.2 [1.0, 1.4] | 1.1 [0.9, 1.3] | 1.3 [1.1, 1.6] |
| **Creatinine (μmol/L)** | 79.0 [66.0, 98.0] | 84.0 [73.0, 104.0] | 67.0 [57.0, 83.0] |
| **HbA1c (mmol/mol)** | 54.0 [48.0, 65.0] | 55.0 [48.0, 65.0] | 54.0 [46.0, 65.0] |
| **Triglycerides (mmol/L)** | 1.8 [1.2, 2.5] | 1.8 [1.2, 2.6] | 1.7 [1.2, 2.3] |
| **Metformin (%)** | 646 (71%) | 424 (70%) | 222 (72%) |
| **DPP-4 inhibitors (%)** | 96 (11%) | 63 (10%) | 33 (11%) |
| **Sulfonylurea (%)** | 144 (16%) | 102 (17%) | 42 (14%) |
| **GLP-1 receptor agonist (%)** | 204 (22%) | 122 (20%) | 82 (27%) |
| **Insulin (%)** | 426 (47%) | 289 (48%) | 137 (44%) |
| **Beta blockers (%)** | 253 (28%) | 186 (31%) | 67 (22%) |
| **ACE inhibitors (%)** | 353 (39%) | 253 (42%) | 100 (32%) |
| **Angiotensin II receptor blockers (%)** | 362 (40%) | 239 (40%) | 123 (40%) |
| **Calcium antagonists (%)** | 298 (33%) | 212 (35%) | 86 (28%) |
| **Diuretics (%)** | 477 (52%) | 322 (53%) | 155 (50%) |
| **Statins (%)** | 714 (78%) | 489 (81%) | 225 (73%) |
| **Aspirin (%)** | 554 (61%) | 386 (64%) | 168 (54%) |

<sup>1</sup>Continuous variables: median [Q1, Q3]. Categorical variables: n (%). Missing rows shown where applicable.

As shown in the present investigation and by others^6,13,19^, the lipid profile reflects the metabolic status of an individual and thus is subjected to factors, such as disease conditions, use of lipid-lowering medication, diet and smoking^19^. Therefore, we additionally investigated the sex-specific circulating lipid profile with further adjustment for relevant parameters (see Methods for details). In general, a high degree of agreement between this and the age-adjusted model assessing lipid-sex associations was observed (Figure 4b) and all lipid (sub)classes remained statistically significant, except HexCer.

For a more detailed view on the lipid differences between men and women, we expanded our analysis to molecular lipid levels, since individual lipid species could be influenced in their biological function due to their acyl-chain and saturation status (Figure 4c). From the analyzed lipid species, 173 lipids from the age-adjusted model showed a statistically significant association with sex (FDR adjusted p-value <0.05). Of those lipids, 42 lipids were from the PC-and 34 from the SM class and all these lipids were higher in women. In line with that, individual lipid species from PC and SM class displayed the strongest associations. For example, the lipid species SM(d18:2/14:0), PC(34:5) and PC(36:6), respectively, were -36.0% (95% CI, - 40.7 to -30.9), -32.4% (95% CI, -40.3 to -23.5) and -32.1% (95% CI, -37.9 to -25.7) lower in men relative to women. Whereas TG(18:1_40:1), CAR(20:4) and CAR(26:0), respectively, were 32.4% (95% CI, 12.2 to 56.2), 31.0% (95% CI, 22.2 to 40.4), and 20.8% (95% CI, 13.9 to 28.2) higher in men.

In summary, our data show a clear sex-specific lipid profile with higher levels of SMs, PCs, PC(P)s, PC(O)s and PEs in women in both models. Thus, our data highlights the need to consider in the analysis and interpretation of lipidomics data sex-specific differences in people living with T2D especially in clinical settings.

## DISCUSSION

Plasma lipid profiling by, for example, LC-MS/MS-based targeted lipidomics, enables the detailed analysis of circulating lipids and thus is a promising tool for clinical studies^24^.

The established LC-MS/MS targeted lipidomics method detects 513 lipid transitions in human plasma belonging to 25 lipid (sub)classes via a dMRM approach with high robustness (mean CV across all lipids: 8.6%) allowing the characterization of an insightful circulating lipid signature in an individual at a given time. While the average platform-specific variability was low, heterogeneity between lipid classes in terms of accuracy was observed. In general, CARs, LPCs and SMs had the lowest technical variability (mean CV in the respective classes <10%) whereas CEs and DG had higher platform-specific variability (23.1% and 17.0%, respectively). The same trend for the lipid class-specific variability has also been reported previously^9,10,19^, and may be attributed to the structural and chemical features of the lipids classes which is important for the analytical procedure, like chromatographic resolution and/or MS transition detectability.

The greatest challenge here was with CEs. Even though they are highly abundant in plasma^25^, only a few CEs could be detected with the instrumental settings used, and those few species were measured with rather high CVs. This is mainly due to the chemical instability of CEs under high energy conditions as found during ionization in the ion source, leading to ion source fragmentation^26,27^. Only CE species with a high degree of unsaturation are stable enough under such conditions to be substantially detected^27^, as exemplified by CE(20:4) and CE(22:5) in our data. This is in line with the observation that for CE lipids the temporal variability is nearly as high as the platform-specific variability. In contrast, PS lipids showed a high temporal variability, but the platform-specific variability of PSs was low. PS lipids are strong anti-inflammatory signals since they function as “eat-me” signals of apoptotic cells and inhibit the production of inflammatory mediators, interleukin (IL)-6 and IL-8^19,28^. Therefore, PS lipids are tightly regulated and have a short half-life^29^, providing a possible explanation for the high variability observed within an individual over time.

Besides inflammation, also diet directly affects the plasma lipid profiles, though mainly TGs and DGs^19,30^. Their breakdown products may subsequently influence CARs and FFAs lipids and thus lipid metabolism is also an important contributor to lipid level variability^19,31^. This notion is supported by our observation that medium-chain lipid showed a trend towards higher temporal variability compared to long- and very-long-chains in our acyl-chain specific analysis. Differences in lipid metabolism are also a main source for the inter-individual lipid level variability^19^. Accordingly, we also found high inter-individual variability with TGs and DGs in our data. Assessing the lipid profile variability within an individual and between individuals in a given population is crucial, since these parameters are important for clinical biomarker development, such as lipid-based ones^14,19^. We observed that all lipid classes exhibited higher between-individual than intra-individual variability, indicating that plasma lipid profiles are specific for a certain condition and can fluctuate due to various individual factors such as environment, disease status or (sex-specific) genetic background^18,19^. Thus, our temporal variability data, mapped across lipid classes and acyl-chain groups, will be important for identifying lipid groups that can be used for patient stratification to enable more precise healthcare, independent of the actual clinical goal.

In addition, we also assessed circulating lipid profiles in 914 men and women living with T2D. Interestingly, 33% of the measured lipids displayed a statistically significant difference in their levels between men and women. This is consistent with previous reports identifying sex as a major driver of inter-individual lipid profile variation^14,19^. We observed higher lipid levels in women for the SM, PC and PE classes, whereas higher levels in men were found for CAR and CAR;O^13,19^. In contrast to lipidomic results from a large Australian prospective study^13^, we found also that the alkyl and alkenyl subclasses of both PE and PC were higher in women in our cohorts. This discrepancy may be explained by the fact that sex can influence the trajectory of diabetes and diabetes associated diseases^32^ and that the prevalence of diabetes in the Australian cohort was only 3.8%. Thus, higher levels of PC(O), PC(P), PE(O) and PC(P) in women might be specific to our T2D cohort.

At the individual lipid species level, we observed substantial overlap in sex associations for SM and PC species compared with published data^13,21,33^. For example, approximately 70% of the sex-associated SM lipid species overlapped with findings from Beyene et al.^13^. This overlap was evident not only qualitatively but also quantitatively, as the lipid species showing the strongest sex associations within the SM lipid class were nearly identical across datasets. Similarly, albeit slightly weaker, concordance was observed for PC species. Both, SMs and PCs are predominately transported in circulation by HDL particles^34^ and since HDL-C is higher in women compared to men in the Thousand&2 cohort, higher SM and PC lipid levels in women were expected. Another contributing factor is the sex-specific enzymatic turnover of lipids. In men, the enzymatic activities of lipoprotein-associated phospholipase A2 (Lp-PLA2), and lecithin cholesterol transferase (LCAT) are higher^13,19^. These enzymes are involved in the hydrolysis of PC lipids to LPC and FFA, or in the transesterification of FAs from PC to cholesterol generating CE and LPC^13,19^. Consequently, PC levels are generally lower and LPC levels are higher in men. Although we did not observe statistically significant higher LPC levels in men in our data, the mean LPC levels were higher, consistent with published reports showing higher LPC levels in men^13^. Together, our results and the existing literature highlight the importance of considering, and where possible applying, sex-stratified analyses in lipid research^6,13,14,19^. Moreover, our sex-dependent plasma lipid profiles are largely consistent with existing literature^13,33,35,36^, while providing additional insights due to the size and T2D-specific nature of our cohorts.

Overall, our data provide valuable insights into lipid level variability in people with T2D by assessing platform-specific, temporal, and inter-individual variability. Through this approach, stable and unstable lipid classes as well as acyl-chain groups were identified. This information will be important for future efforts in MS-based lipidomic biomarker development in the context of clinical T2D management. Our large T2D-specific cohort demonstrates that sex-specific differences must be considered in diabetes-related lipid research. In future, it will be valuable to investigate lipid profiles in individuals living with diabetes-associated comorbidities to identify lipid biomarkers and elucidate lipid-mediated molecular mechanisms underlying these diseases.

### Strength and limitations

One limitation of this study is the small sample size for determining temporal lipid variability. Moreover, the accurate determination of the contribution of the technical variability to the temporal lipid level fluctuation within an individual requires repeated measurements of each individual sample. However, the platform-specific variability was based on repeated measurements of a pooled plasma sample and thus may not fully represent the technical variability in temporal lipid variations. While the lipidomics data captured lipid signal for a wide range of lipid species and (sub)classes, some lipid features remain underrepresented due to a low number of accurately measured lipid species, which may compromise the holistic view of these lipid classes.

Key strengths of this study are the use of well-defined and homogenous cohorts and the large sample size for the determination of sex-specific lipid levels. Moreover, the detailed characterization of the lipidomic analysis allowed us account for methodical biases, as e.g. with CE.

## METHODS

### The LIRAFLAME Cohort

In this study, we analyzed plasma samples of three time points from the placebo arm (N=51) of the LIRAFLAME randomized controlled trial^37^. The trial was conducted between 2017-19 at the Steno Diabetes Center Copenhagen. Here people with type 2 diabetes were included in a randomized, double-blind, placebo-controlled, parallel-group trial. The inclusion criteria were as follows: age >50 years; HbA1c ≥ 48 mmol/mol (6.5%); estimated glomerular filtration rate ≥30 mL/min/1.73 m2 (estimated by Chronic Kidney Disease Epidemiology Collaboration (CKD-EPI) formula); stable glucose-lowering and cholesterol-lowering treatment (minimum 4 weeks). Key exclusion criteria were: type 1 diabetes; chronic or previous acute pancreatitis; other treatment which in the investigator’s opinion could interfere with the study.

This study was carried out in concordance with the principles of the Declaration of Helsinki and ethics approval was granted by local ethics committee (H-16044546) and the Danish Medicines Agency (2016110109). Participants provided written informed consent before being included.

### The Thousand&2 Cohort

The Thousand & 2 Study is a representative sample of patients with type 2 diabetes initiated in October 2011 from Steno Diabetes Center (SDC) and Center for Diabetes Research (CfD), Department of Medicine, Herlev and Gentofte Hospital, University of Copenhagen^23^. Patients from the primary sector with inadequate glycaemic control or diabetes-related complications are referred to the centers to optimize medical treatment. All patients with type 2 diabetes followed at either center were eligible to participate and were recruited at SDC or CfD^23^. The study was conducted in accordance with the Helsinki Declaration, approved by The Danish National Committee on Biomedical Research Ethics, amendment to protocol no. H-3-2009-139. All participants gave written informed consent.

### Lipid Extraction

Lipid extraction was performed as described previously^5,9^. In brief, 10 µL of plasma was mixed with 100 µL of butanol:methanol (1:1) containing 10mM ammonium formate and a mixture of 21 stable isotope-labelled internal lipid standards spanning the entire panel of measured lipid classes (Supplementary). Samples were vortexed thoroughly and set in a sonicator bath for 15 mins maintained at room temperature and subsequently incubated for 45 min with 1000 rpm at 4°C. Samples were then centrifuged (14,000xg,10 min, 4°C) before transferring the into MS vials with glass inserts for analysis. Sample have been stored at -80°C till analysis.

### Liquid Chromatography Mass Spectrometry

Analysis of plasma extracts was performed on an Agilent 6460C QQQ mass spectrometer with an Agilent 1290 series HPLC system and a ACQUITY UPLC BEH C18 Column (2.1x100mm 1.7 µm, Waters) with the thermostat set at 45°C. Mass spectrometry analysis was performed by positive/negative ion mode switching with dynamic scheduled multiple reaction monitoring (dMRM). To determine the specific dMRM measurement windows for each lipid analyte, we used pooled plasma samples from each cohorts analyzed. Mass spectrometry settings and MRM transitions for each lipid class, subclass and individual species are shown in the Supplementary Information.

The LC buffer consisted of solvent A) 50% H2O / 30% acetonitrile / 20% isopropanol (v/v/v) containing 10mM ammonium formate and 2 mM ammonium fluoride. Solvent B consisted of 1% H2O / 9% acetonitrile / 90% isopropanol (v/v/v) containing 10mM ammonium formate and 2 mM ammonium fluoride. We utilized a stepped linear gradient with a 16-minute cycle time per sample and a 5 µL sample injection. The gradient detail can be found in the Supplementary Information.

The following mass spectrometer conditions were used; gas temperature, 325°C, gas flow rate 9 L/min, nebulizer 35 psi, Sheath gas temperature 350°C, capillary voltage 3000V (both positive and negative) and sheath gas flow 11 L/min. Isolation widths for Q1 and Q3 were set to ‘‘unit’’ resolution (0.7 amu).

### LC-MS/MS Data analysis and lipid quantification

Chromatographic peaks were integrated using the Mass Hunter (B.07.00, Agilent Technologies) software and assigned to a specific lipid species based on MRM (precursor/product) ion pairs and their chromatographic behavior. To quantify lipid analytes and internal standards, peak area of each analyte was used, resulting in relative quantification of lipid species in each sample. Before statistical analysis, lipid species with missing values >30% over the entire dataset and outliers deemed to be of technical origin were excluded from downstream analysis. Subsequently, peak area of each analyte was normalized to the peak area of the corresponding internal standard. Batch effects during the lipidomic analysis of Thousind&2 were corrected using a median centering approach utilizing QC samples. Quantification of lipid classes was determined as the sum composition of the lipid species within each class.

### Linearity Experiments

To assess linearity of response for a representative individual lipids for each lipid classes, titration experiments were performed. Following lipids have been used: CAR(16:0)d3, CE(18:1)d7, Cer(d18:1-d7/16:0), Cer(d18:1-d7/18:0), Cer(d18:1-d7/24:0), Cer(d18:1-d7/24:1), DG(15:0-18:1)d7, FA(18:0)d35, LPC(18:1)d7, LPE(18:1)d7, PA(15:0_18:1)d7, PC(15:0_18:1)d7, PC(P-18:0/18:1)d9, PE(15:0_18:1)d7, PG(15:0_18:1)d7, PI(15:0_18:1)d7, PS(15:0_18:1)d7, SM(d18:1/18:1)d9, TG(15:0_33:1)d7 and purchased form Avanti Polar Lipids except FA(18:0)d35 and CAR(16:0)d3 which have been purchased from Cayman Chemical Company. Various lipids concentration were spiked into extracted plasma samples and analyzed. Response was determined from the areas of the tested lipid species and plotted. To assess the linearity an R^2^ value was calculated for each lipid.

### Statistical analysis

All data analyses were done in R.

In the LIRAFLAME Cohort, between-individual variability and within-individual variability were assessed through cross-sectional and longitudinal relative standard deviations (RSD), and respectively, their ratio. Cross-sectional RSD, estimating the between-individual variability, was calculated across individuals at each time point, and averaged over the time points. Longitudinal RSD, estimating the within-individual variability over time, was calculated across the three time points for each individual, and averaged over the individuals.

The cross-sectional analysis of associations between sex and lipids/lipid (sub)classes in the Thousand&2 cohort was performed using multiple linear regression (lm()) in two steps; one model included age + sex as predictors and the second model included age + sex + diabetes duration (years) + BMI (kg/m2) + HbA1c (mmol/mol) + smoking (never, former, current) + treatment with DPP4 inhibitor, a GLP-1 recepter agonist, metformin, sulphonyl urea, statins (all: yes, no). The lipid data was log10 transformed for analysis and backtransformed and converted to percent for presentation.

## Data availability

In line with the current regulation of General Data Protection Regulation (https://gdpr-info.eu/) to maintain patient confidentiality, individual-level clinical and lipidomics data generated in this study cannot be made publicly available. Access to the data can be granted by contacting the lead authors with additional approval from the Danish Data Protection Agency and the ethics committee.

## ACKNOWLEDGEMENTS

The authors would like to thank all participants and acknowledge the work of study nurses from Steno Diabetes Center Copenhagen, and Center for Diabetes Research (CfD), Department of Medicine, Herlev and Gentofte Hospital, Denmark. A special thanks to Annette Frost Bjerre for technical assistance with sample preparation for lipidomic sample analysis.

## AUTHOR CONTRIBUTIONS

SMK, TS, MBB, KS and CLQ contributed to study design and data interpretation. SMK performed targeted lipidomics sample analysis. SMK processed and analyzed the lipidomics data, together with MBB and TS. SMK and CLQ interpreted the lipidomics data. MBB performed statistical analysis of the clinical data. SMK drafted the manuscript, and the final version was critically reviewed and approved by all authors.

## Supplementary Figures

**Sup Fig. 1:**
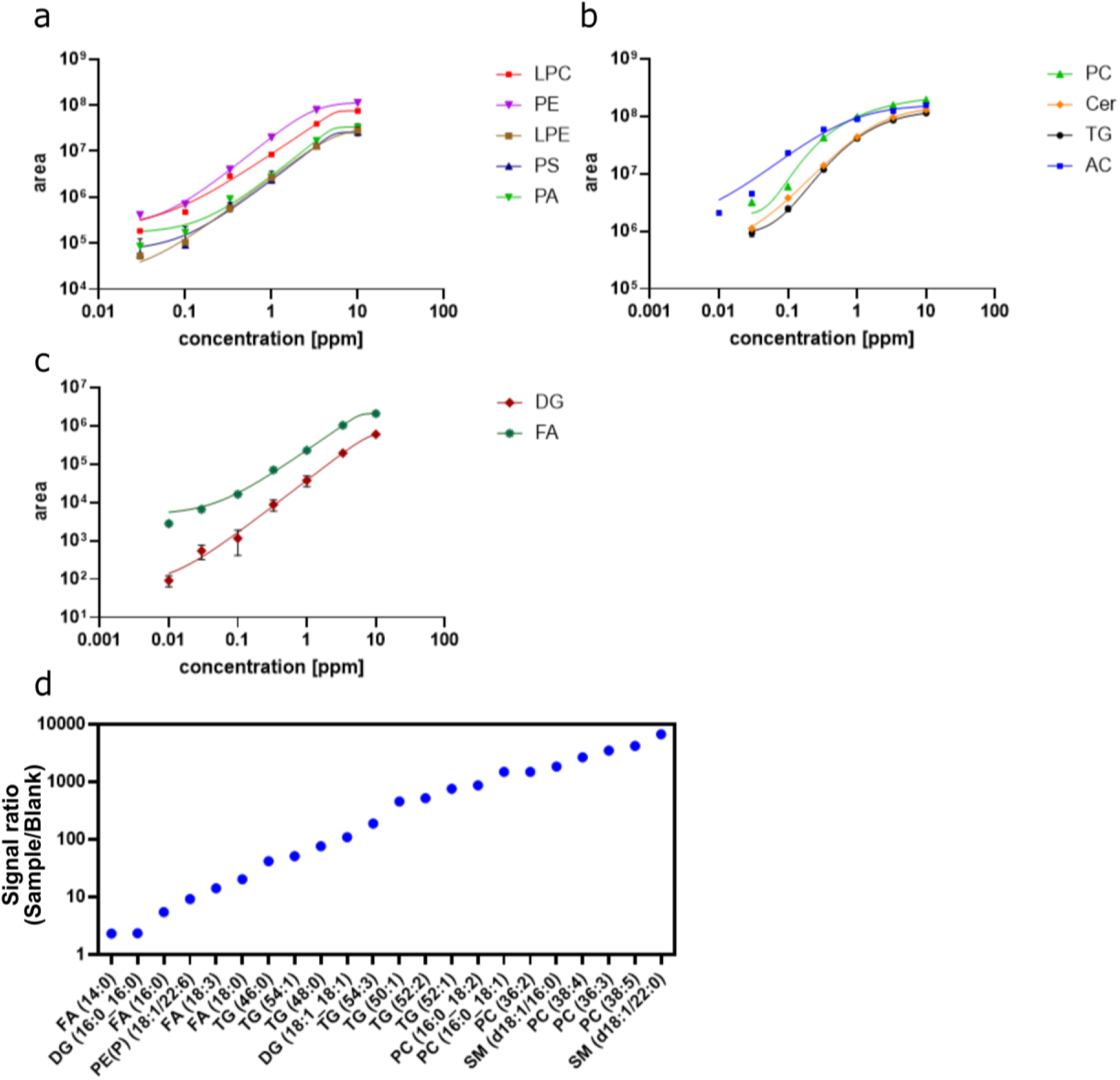
Linearity and carry-over effects of measurements. **a-c:** A linear increase range in lipid signal is observed for studied lipid. Labelled lipids were spiked into an extracted plasma of quality control samples at their respective concentrations to generate response curves. Signal saturation and limit of detection were reached with all lipids. **d:** Estimate of the carry-over effect, calculated by the ratio of the signal intensity in a plasma quality control sample (sample) through a blank sample, for the most affected lipid species.

**Table S1:** Clinical characteristics at baseline of individuals for the longitudinal data. Data for continuous variables are presented as median [IQR] and categorical variables as number of individuals and as percentage in brackets. HbA1c: hemoglobin A1c, HDL: high-density lipoprotein, LDL: low-density lipoprotein.

| Individuals | n = 51 |
| --- | --- |
| **Sex (women)** | 10 [19.6%] |
| **Age (years)** | 66.9 [7.8] |
| **Diabetes duration (years)** | 10.2 [5.7-19.2] |
| **Body mass index (kg/m<sup>2</sup>)** | 29.3 [3.8] |
| **Systolic blood pressure (mmHg)** | 137.3 [19.7] |
| **Diastolic blood pressure (mmHg)** | 78.9 [8.4] |
| **Smoking current** | 4 [7.8] |
| **Urinary albumin creatinine ratio, mg/g** | 6.0 [3.5–14.5] |
| **LDL cholesterol (mmol/L)** | 2.1 [0.69] |
| **HDL cholesterol (mmol/L)** | 1.3 [0.3] |
| **HbA1c (mmol/mol)** | 58.0 [10.6] |
| **Triglycerides (mmol/L)** | 1.6 [0.8] |
| **Lipid-lowering treatment** | 42 [82.4%] |

**Sup Fig. 2:**
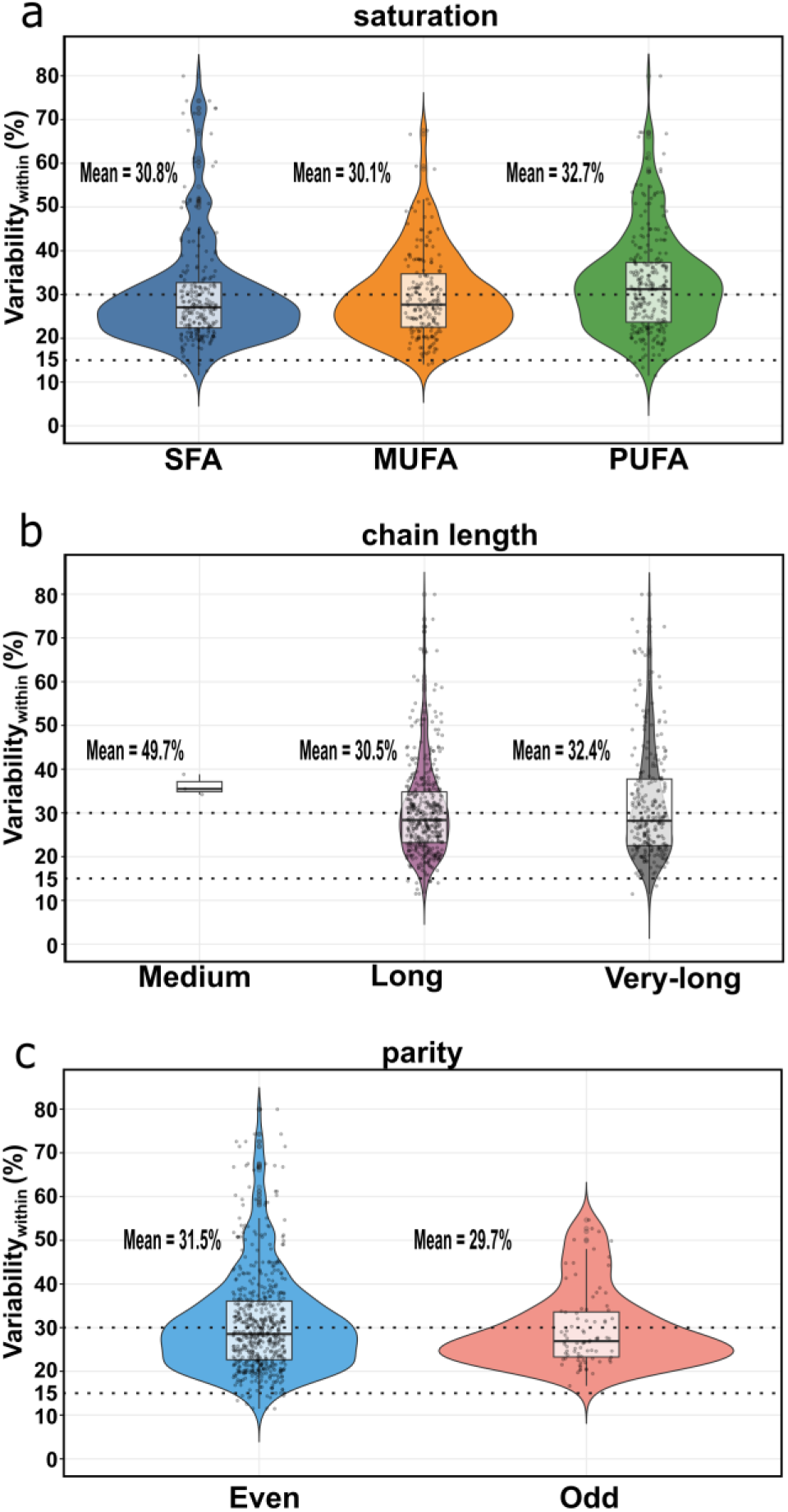
Temporal variability of acyl groups. Within-individual variability of lipid acyl groups is comparable between each group. Mean variability in percent (%) is shown for each lipid acyl groups. Box plots display the distribution of variability estimates across detected lipid species within each acyl groups. Dotted lines indicate 15% and 30% variability. Acyl-chains are grouped by acyl-chain saturation (a) (SFA, saturated fatty acid; MUFA, mono-unsaturated fatty acid; PUFA, poly-unsaturated fatty acid) by acyl-chain length (b) (medium-(C6-12), long-(C13-21) and very-long chain-lengths (>C21)), and by parity (c) of the carbon atoms number in the acyl chain. Data table with values for each lipid species is provided in supplemental source Table 5.

**Sup Fig. 3:**
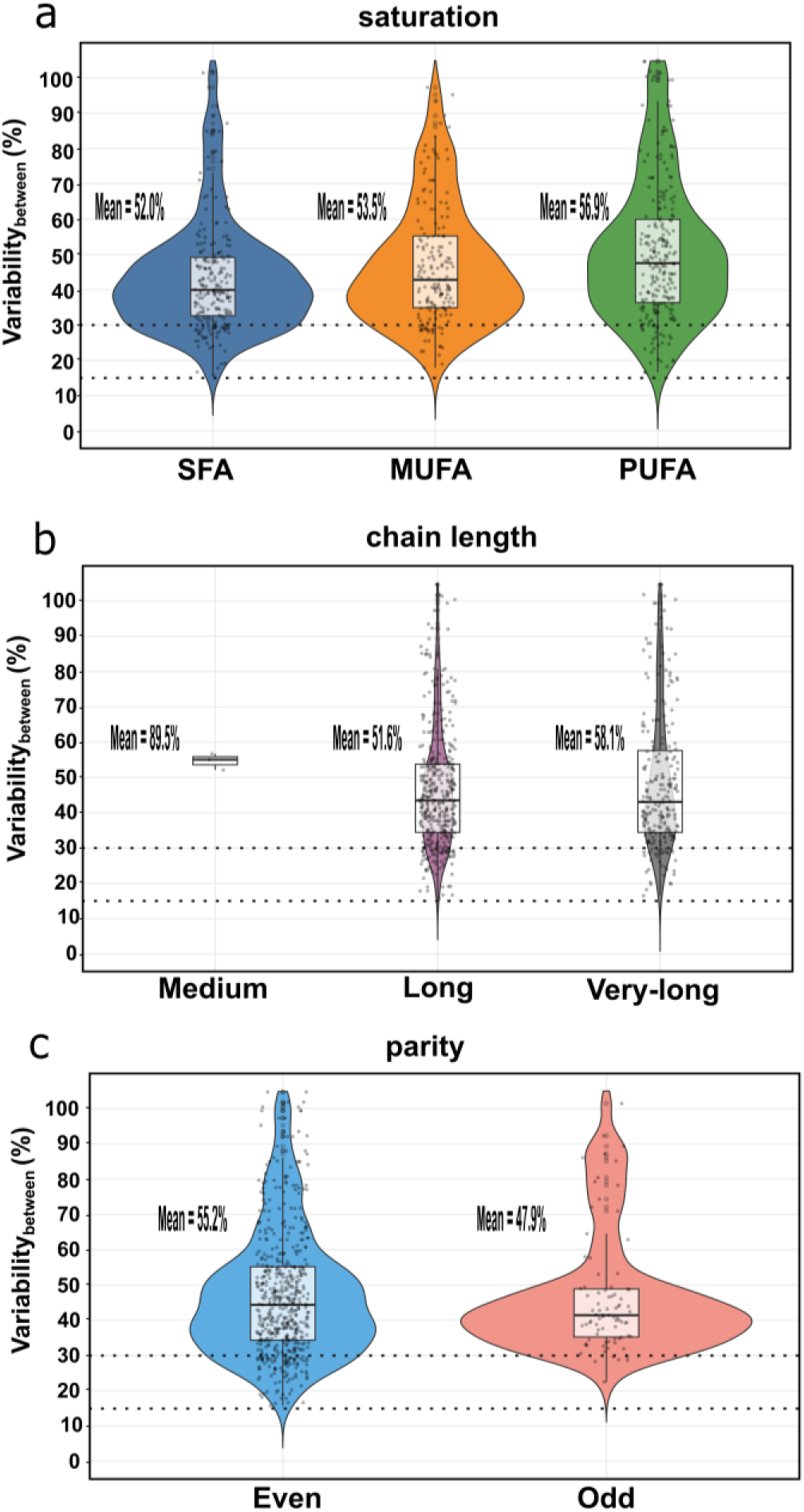
Inter-individual variability of acyl groups. Inter-individual variability of lipid acyl groups is comparable between each group. Mean variability in percent (%) is shown for each lipid acyl groups. Box plots display the distribution of variability estimates across detected lipid species within each acyl groups. Dotted lines indicate 15% and 30% variability. Acyl-chains are grouped by acyl-chain saturation (a) (SFA, saturated fatty acid; MUFA, mono-unsaturated fatty acid; PUFA, poly-unsaturated fatty acid) by acyl-chain length (b) (medium-(C6-12), long-(C13-21) and very-long chain-lengths (>C21)), and by parity (c) of the carbon atoms number in the acyl chain. Data table with values for each lipid species is provided in supplemental source Table 5.

**Table S2:** Lipid class levels by sex of lipidomics data from the Thousand&2 cohort. . The median and interquartile range of lipid (sub)class levels is shown for men, women and combined (All).

| Lipid class | All<br>N = 914<sup>1</sup> | Men<br>N = 605<sup>1</sup> | Women<br>N = 309<sup>1</sup> | p-value<sup>2</sup> |
| --- | --- | --- | --- | --- |
| Acylcarnitine (CAR) | 20.9 [15.7, 26.8] | 21.2 [16.1, 27.1] | 19.8 [15.2, 26.3] | 0.045 |
| Hydroxylated acylcarnitine (CAR;O) | 1.0 [0.7, 1.4] | 1.0 [0.8, 1.4] | 1.0 [0.7, 1.3] | 0.084 |
| Cholesterylester (CE) | 1.4 [0.9, 2.2] | 1.3 [0.8, 2.1] | 1.5 [1.0, 2.3] | 0.001 |
| Sphingomyelin (SM) | 111.8 [90.8, 131.5] | 104.9 [87.5, 123.2] | 125.5 [103.5, 145.7] | <0.001 |
| Ceramide (Cer(d)) | 5.8 [4.4, 7.3] | 5.7 [4.4, 7.1] | 5.8 [4.4, 7.7] | 0.2 |
| Deoxyceramide (Cer(m)) | 2.2 [1.5, 3.4] | 2.3 [1.5, 3.4] | 2.2 [1.4, 3.4] | 0.2 |
| Dihexosylceramide (Hex2Cer) | 1.1 [0.9, 1.4] | 1.1 [0.9, 1.4] | 1.1 [0.9, 1.4] | >0.9 |
| Monohexosylceramide (HexCer) | 0.7 [0.5, 0.9] | 0.7 [0.5, 0.8] | 0.7 [0.5, 0.9] | 0.030 |
| Trihexosylceramide (Hex3Cer) | 0.5 [0.3, 0.6] | 0.4 [0.3, 0.6] | 0.5 [0.4, 0.6] | 0.002 |
| Triacylglycerol (TG) | 312.0 [195.5, 480.8] | 315.1 [200.3, 487.7] | 307.6 [189.8, 455.2] | 0.4 |
| Diacylglycerol (DG) | 119.8 [84.1, 172.2] | 118.2 [84.7, 174.7] | 123.0 [84.0, 162.4] | >0.9 |
| Free fatty acids (FFA) | 63.5 [44.9, 86.6] | 61.0 [43.6, 83.6] | 68.1 [48.7, 95.6] | 0.004 |
| Lysophosphatidylcholine (LPC) | 145.3 [116.9, 177.6] | 145.1 [118.5, 179.3] | 145.6 [113.9, 176.0] | 0.5 |
| Lysoalkylphosphatidylcholine LPC(O) | 3.7 [2.8, 4.7] | 3.8 [2.8, 4.8] | 3.6 [2.7, 4.6] | 0.054 |
| Lysoalkenylphosphatidylcholine LPC(P) | 0.6 [0.5, 0.8] | 0.7 [0.5, 0.8] | 0.6 [0.5, 0.8] | 0.2 |
| Lysophosphatidylethanolamine (LPE) | 8.5 [6.4, 11.2] | 8.2 [6.3, 11.2] | 8.9 [6.8, 11.2] | 0.054 |
| Lysoalkenylphosphatidylethanolamine LPE(P) | 0.5 [0.3, 0.6] | 0.5 [0.3, 0.6] | 0.4 [0.3, 0.6] | 0.9 |
| Phosphatidic acid (PA) | 0.00 [0.00, 0.00] | 0.00 [0.00, 0.00] | 0.00 [0.00, 0.00] | 0.3 |
| Phosphatidylcholine (PC) | 458.0 [372.4, 540.2] | 433.2 [364.9, 514.0] | 499.6 [418.0, 588.7] | <0.001 |
| Alkylphosphatidylcholine PC(O) | 59.1 [47.8, 71.3] | 57.6 [47.0, 68.9] | 63.4 [51.1, 75.4] | <0.001 |
| Alkenylphosphatidylcholine PC(P) | 33.7 [26.7, 41.4] | 32.8 [25.6, 39.7] | 36.6 [28.4, 44.5] | <0.001 |
| Phosphatidylethanolamine (PE) | 41.1 [29.3, 55.5] | 39.1 [27.8, 52.8] | 45.8 [32.8, 59.8] | <0.001 |
| Alkylphosphatidylethanolamine PE(O) | 3.5 [2.7, 4.6] | 3.4 [2.6, 4.5] | 3.6 [2.9, 4.8] | 0.007 |
| Alkenylphosphatidylethanolamine PE(P) | 38.2 [30.0, 48.2] | 37.7 [29.5, 47.3] | 39.1 [31.2, 49.9] | 0.024 |
| Phosphatidylserine (PS) | 0.01 [0.01, 0.03] | 0.01 [0.01, 0.03] | 0.01 [0.01, 0.02] | >0.9 |
| Total lipid sum | 1,493.6 [1,217.3, 1,807.2] | 1,456.0 [1,188.8, 1,772.7] | 1,549.6 [1,269.0, 1,875.1] | 0.009 |

<sup>1</sup>Median [Q1, Q3]
<sup>2</sup>Wilcoxon rank sum test

**Sup Fig. 4:**
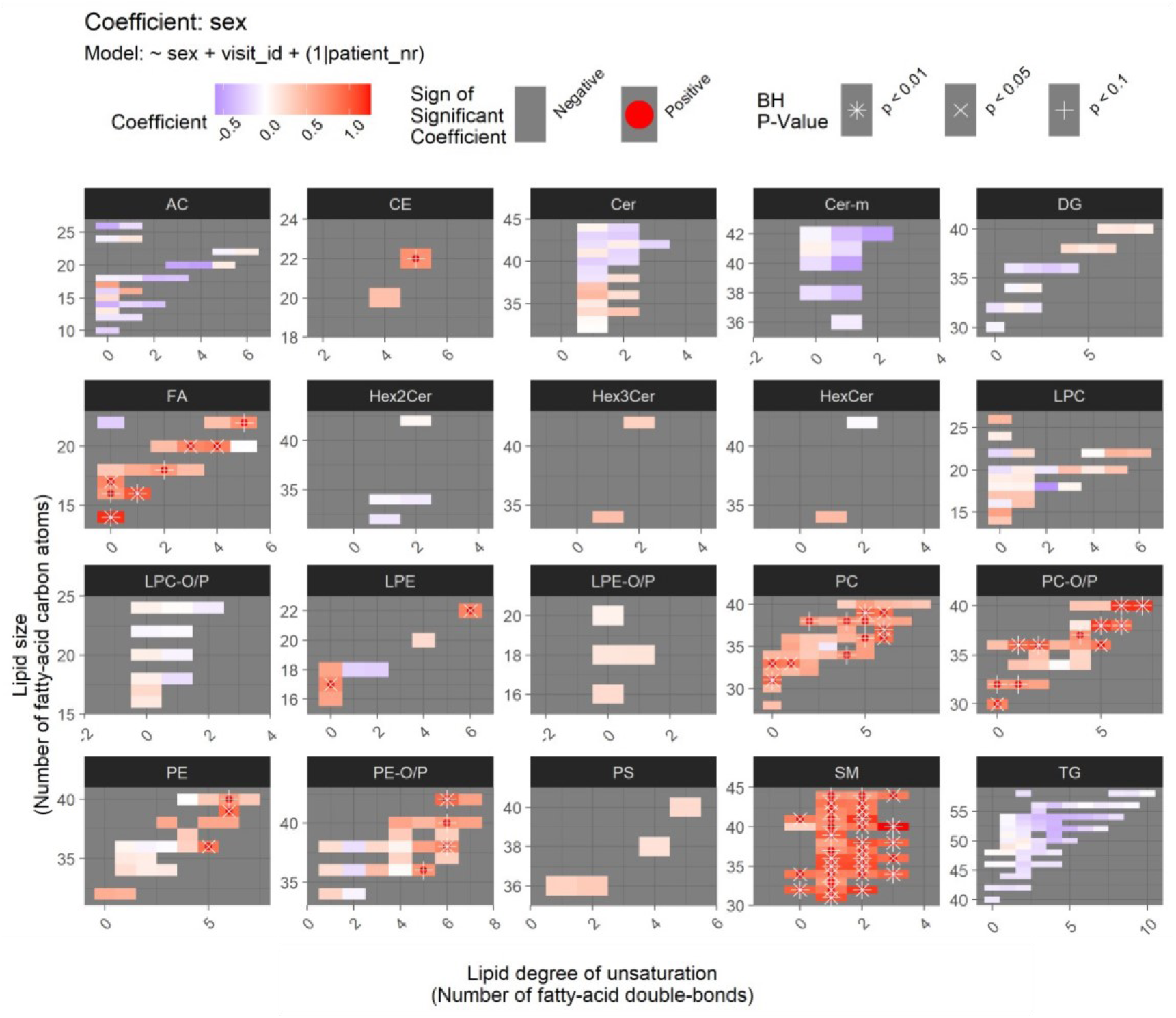
Difference in the lipidome between men and women for the longitudinal data. Lipids are grouped by their class into panels within each panel, each colored rectangle corresponds to one lipid species, and its location in the x-axis and y-axis, respectively, shows its size (number of carbon atoms in the fatty acid chains) and its level of unsaturation (number of double bonds). Blue, red and white rectangles, respectively, indicate lower, higher and same levels in women compared to men (based on the regression coefficient from the linear regression model). Statistical significance of the difference is annotated by the symbols “*,” “x” and “+,” respectively, corresponding to p < 0.01, 0.05 and 0.1.

